# Seroclearance of HBsAg in Chronic Hepatitis B Patients After a Tolerance Breaking Immuno-Therapy: GM-CSF, followed by A Recombinant HBV Vaccine

**DOI:** 10.1101/2022.04.18.22273874

**Authors:** Shuang Geng, Feifei Yang, Hongyu Jia, Gan Zhao, Weidong Zhao, Jie Yu, Haoxiang Zhu, Huan Cai, Lishan Yang, Shuren Zhang, Xian Zhou, Chaofan Li, Fang Yu, Xiang Jin, Shijie Zhang, Xianzheng Wang, Yida Yang, Jimin Zhang, Bin Wang

## Abstract

**Background:** Chronic hepatitis B (CHB) remains incurable due to the immune system’s tolerance towards the hepatitis B virus (HBV) surface antigen (HBsAg). This study aimed to achieve a functional cure by breaking HBV tolerance through immunotherapy.

**Methods:** CHB patients were treated with either standard nucleotide analog (NA) therapy (Adefovir Dipivoxil, ADV) (cohort 1) or ADV combined with interferon-alpha (IFN-alpha) (cohort 2). Additionally, a third cohort received the THRIL-GM-Vac regimen: three low-dose GM-CSF injections followed by one dose of the HBV vaccine, alongside standard treatment.

**Results:** THRIL-GM-Vac treatment (cohort 3) achieved a significant 2log10 reduction in HBsAg levels in 21.7% of participants compared to 0% and 4.17% in cohorts 1 and 2, respectively. Furthermore, THRIL-GM-Vac significantly reduced HBV-specific tolerogenic T cells (Tregs), explaining the sustained HBsAg decrease. Upregulation of anti-HBV T cell responses confirmed THRIL-GM-Vac’s ability to disrupt HBV tolerance and enhance HBsAg-specific cellular immunity. This suggests its potential effectiveness in treating individuals with moderate to low HBsAg levels.

**Conclusion:** THRIL-GM-Vac treatment in cohort 3 resulted in 8.7% HBsAg clearance alongside Treg depletion and enhanced anti-viral T cell responses. These findings present a promising strategy to overcome immunotolerance and potentially combat chronic HBV infection.

**Significance of This Study:** *What Is Already Known:*

- Persistent viral replication in chronic HBV infection increases the risk of disease progression. Achieving virological suppression is crucial, yet patients with HBsAg still face adverse outcomes, like hepatocellular carcinoma (HCC).
- The ideal treatment goal is a functional cure, or HBsAg loss, which significantly improves clinical outcomes.
- Current treatments include Nucleos(t)ide analogs (NAs) and Interferon (IFNs), with NAs being potent in viral replication inhibition but less effective in HBsAg clearance. IFNs offer a modestly better HBsAg loss rate.
- Combining NAs with IFNs or switching to IFNs has shown some improvement in HBsAg seroclearance in clinical trials.

*New Findings:*

- Previous studies highlighted GM-CSF as potential vaccine adjuvants that boost antitumor and antiviral immunity. Our study demonstrates that a combination of GM-CSF therapy and HBV vaccination (THRIL-GM-Vac), along with ADV and IFN-α as standard treatment, significantly reduces HBsAg levels and enhances anti-HBsAg cell-mediated immunity compared to the standard treatments.
- Specifics include a considerable decrease of HBsAg in 43.5% of patients, with 21.7% exhibiting a major reduction, including HBsAg seroclearance in 8.7% of participants. This response coincided with an increase in cellular immunity markers.

*Clinical Implications:*

- The THRIL-GM-Vac strategy, when combined with conventional antiviral treatments, opens avenues for achieving a functional HBV cure in a more significant proportion of patients.
- Our findings suggest that targeting Treg-dependent immunotolerance correlates with HBsAg reduction, providing a potential immune-surrogate endpoint to predict treatment efficacy.
- This approach offers a promising direction for future research and treatment strategies to meet unmet medical needs in chronic HBV treatment.

## INTRODUCTION

Clearance of serum hepatitis B surface antigen (HBsAg) is the prevalent definition of an HBV infection’s functional cure [1, 2]. However, in persistently infected individuals, immunological tolerance generated by viral persistence remains a formidable obstacle [3, 4].

Establishing immunological tolerance requires modifying regulatory mechanisms, such as T cell exhaustion and the activation of suppressive regulatory cells [4–6]. In particular, Tregs are recognized for their dominance over effector T cells such as CD4+ T helper cells and CD8+ cytotoxic T cells (Ref), and for allowing HBsAg expression in HBV-infected hepatocytes to remain unregulated [7, 8]. Destruction of Tregs is the key to breaking immunological tolerance; hence, it must be prioritized if eradication of serum HBsAg is desired.

Granulocyte-macrophage colony-stimulating factor, or GM-CSF, was one of the first immune-mobilizing drugs found [9, 10] and has been considered a possible adjuvant for anti-viral vaccines, especially HBV vaccines [11–13], for a long time. However, there have been persistent reports of problems surrounding its administration, including uneven immunomodulatory effects and in some cases immunological suppression [14–17]. However, our recent work has been able to harness GM-CSF’s maximum immune-boosting capability through dosing and schedule optimization [18]. In this optimized regimen we call the THRIL-GM-Vac regimen (THRee Injections of Low-dose GM-CSF with one dose of Human HBV Vaccine) [18], we were able to establish substantial levels of HBsAg clearance and anti-HBsAg antibodies in transgenic and AAV-HBV-infected mouse models. Moreover, THRIL-GM-Vac consistently improved the CD8 T cell-mediated immune response. Significant increases in HBsAg-specific IFN-producing CD8 T cells and reductions in Treg cells led to the elimination of tolerance [19, 20]. Here in this clinical research, the translational value of the THRIL-GM-Vac regimen was evaluated to conquer HBsAg clearance in chronic carriers.

## RESULTS

### Patients Characteristics

There was a total of 72 CHB patients who were included in the trial and randomly allocated to one of three groups: 25 patients were treated with ADV (Group 1), 24 patients treated with ADV+IFN-α (Group 2), and 23 patients treated with ADV+IFN-α+THRIL-GM-vac (Group 3). In group 3, two patients discontinued the trial due to adverse events (one with thyroid dysfunction at week 40 and one with rash at week 24). The patient’s baseline characteristics are presented in Table 1.

**Table 1.** The baseline characteristics of the patients.

| **Baseline Characteristics** | **ADV**<br>**(n=25)** | **ADV+IFNα**<br>**(n=24)** | **ADV+IFNα+T**<br>**HRIL-GM-vac**<br>**(n=23)** | **Total**<br>**(n=72)** | **p value** |
| --- | --- | --- | --- | --- | --- |
| **Males n (%)** | **17 (68.0)** | **22 (95.8)** | **20 (87.0)** | **60 (83.3)** | **0.0659** |
| **Age (years)** | **44.0**<br>**(39.0-55.0)** | **40.0**<br>**(32.0-47.5)** | **39.0**<br>**(30.0-50.0)** | **42.0**<br>**(33.0-52.0)** | **0.111** |
| **HBsAg**<br>**(lgIU/mL)** | **2.99**<br>**(2.48-3.27)** | **2.79**<br>**(1.99-3.17)** | **2.64**<br>**(1.78-2.97)** | **2.78**<br>**(2.11-3.19)** | **0.0647** |
| **ALT (IU/L)** | **22.0**<br>**(18.0-29.0)** | **28.0**<br>**(21.0-41.0)** | **26.0**<br>**(19.0-46.0)** | **26.0**<br>**(20.0-35.0)** | **0.180** |
| **AST (IU/L)** | **22.0**<br>**(18.0-28.0)** | **20.0**<br>**(18.5-24.0)** | **22.0**<br>**(19.0-25.0)** | **21.0**<br>**(19.0-25.0)** | **0.884** |
| **WBC**<br>**(\*10<sup>9</sup>/L)** | **5.3**<br>**(4.5 - 5.8)** | **5.5**<br>**(4.9-6.7)** | **5.3**<br>**(4.7-6.6)** | **5.3**<br>**(4.8-6.4)** | **0.628** |
| **HLA-A2,**<br>**Positive (%)** | **14 (56.0)** | **12 (50.00)** | **12 (52.2)** | **38 (52.8)** | **0.913** |

**Notes :**
1. Values are expressed as median (IQR) or the number of patients (%). 2. HBsAg: Hepatitis B surface antigen; ALT: alanine-aminotransferase. AST: aspartate-aminotransferase; WBC: white blood cell; HLA: human leukocyte antigens.

### Enrollment for Each Group

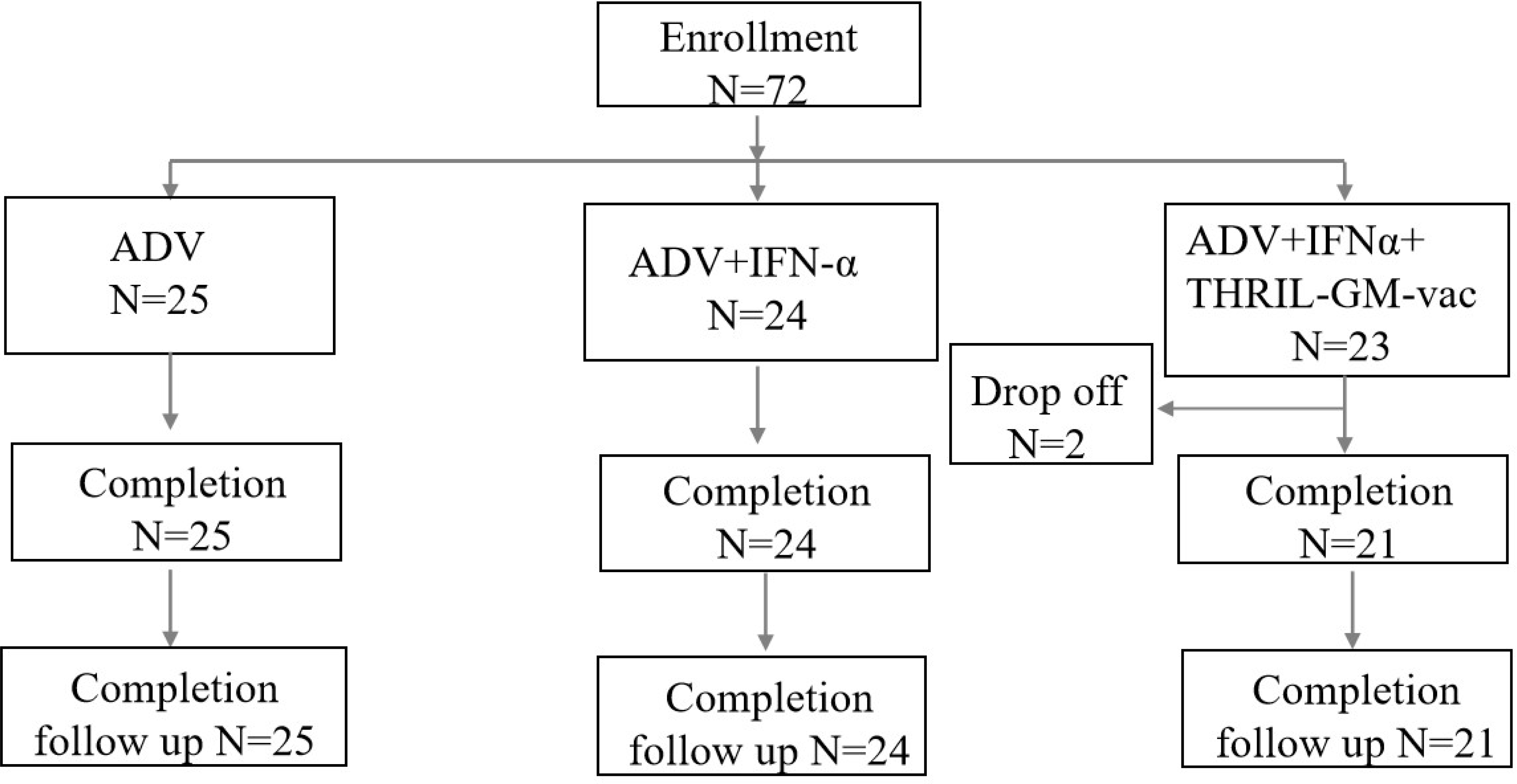

### Assessment of the HBsAg seroclearance under treatment with the THRIL-GM-Vac in addition to ADV and IFN-*α*

Clearance of serum HBsAg was monitored as the most important factor in determining whether or not a functional cure had been achieved, and this approach was carried out per the scheme shown in Figure 1. Patients in Group 1 were given an oral dosage of adefovir NA (ADV) daily, whereas patients in Group 2 were given an oral dose of ADV plus an injection of IFN-α (ADV+IFN-α) daily. Both of these treatments were administered by a protocol known as standard of care (SOC). ADV+IFN-α plus THRIL-GM-vac was the therapy that was administered to participants in Group 3, which was an add-on protocol to the SOC that used the immunization schedule that was discussed before.

**Figure 1.**
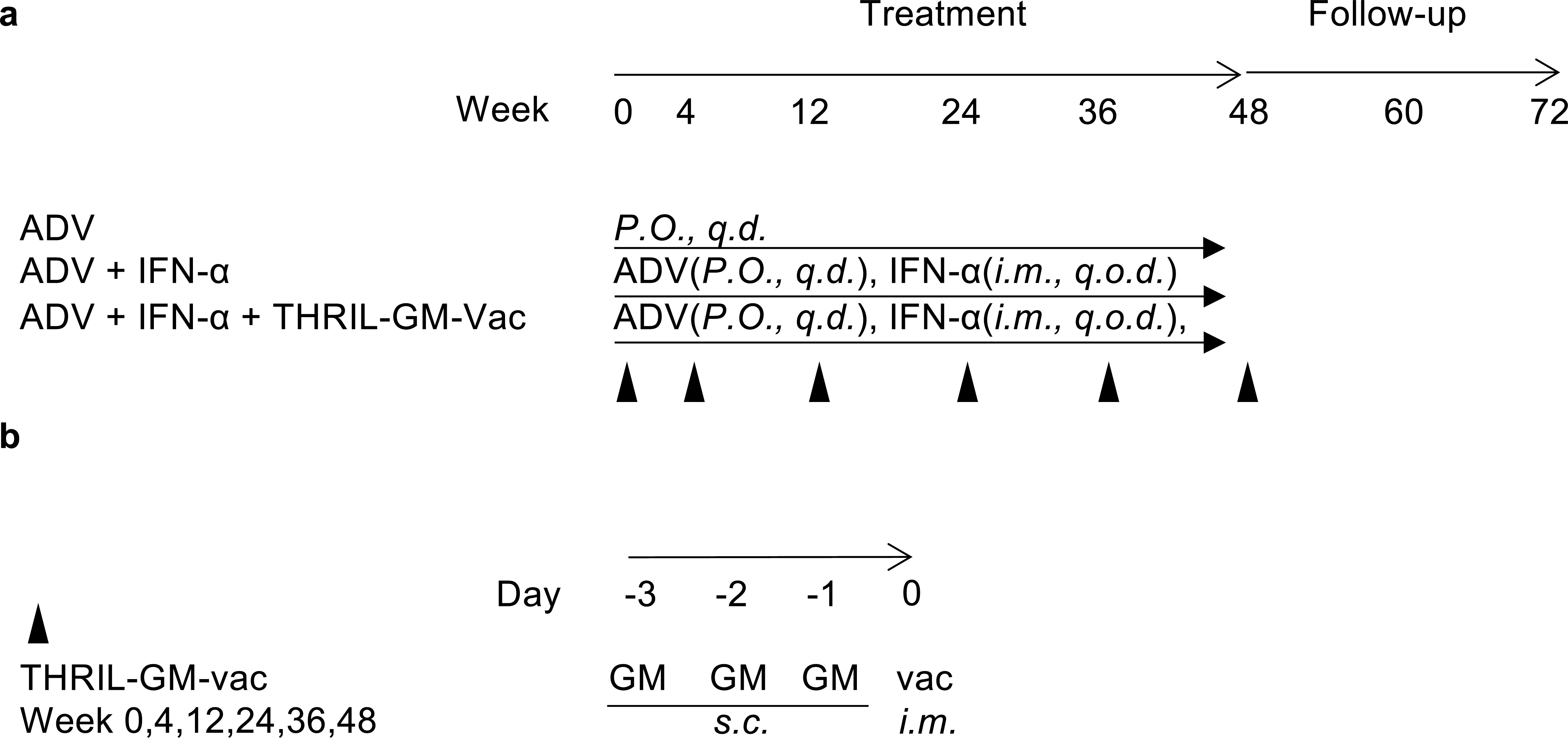
Trial design and scheme. a) Trial schedule and treatment routes in 3 different groups, orally every day (P.O., q.d.), injection intramuscularly injection on alternate days (i.m., qod); b) injections of GM-CSF subcutaneous (s.c.) for three consecutive days followed by an i.m. injection with Human HBV Vaccine at the same site on the 4^th^ day on weeks 0, 4, 12, 24, 36, and 48 as indicated by the solid triangle in (a and b).

Patients in Group 1 did not achieve HBsAg seroclearance at any point throughout their treatment. Seroclearance was seen in one patient (4.17%) in Group 2 who had a low HBsAg baseline at week 24, and it was observed in two patients (8.7%) in Group 3 at week 24 and week 36, respectively, as shown in Figure 2a.

**Figure 2.**
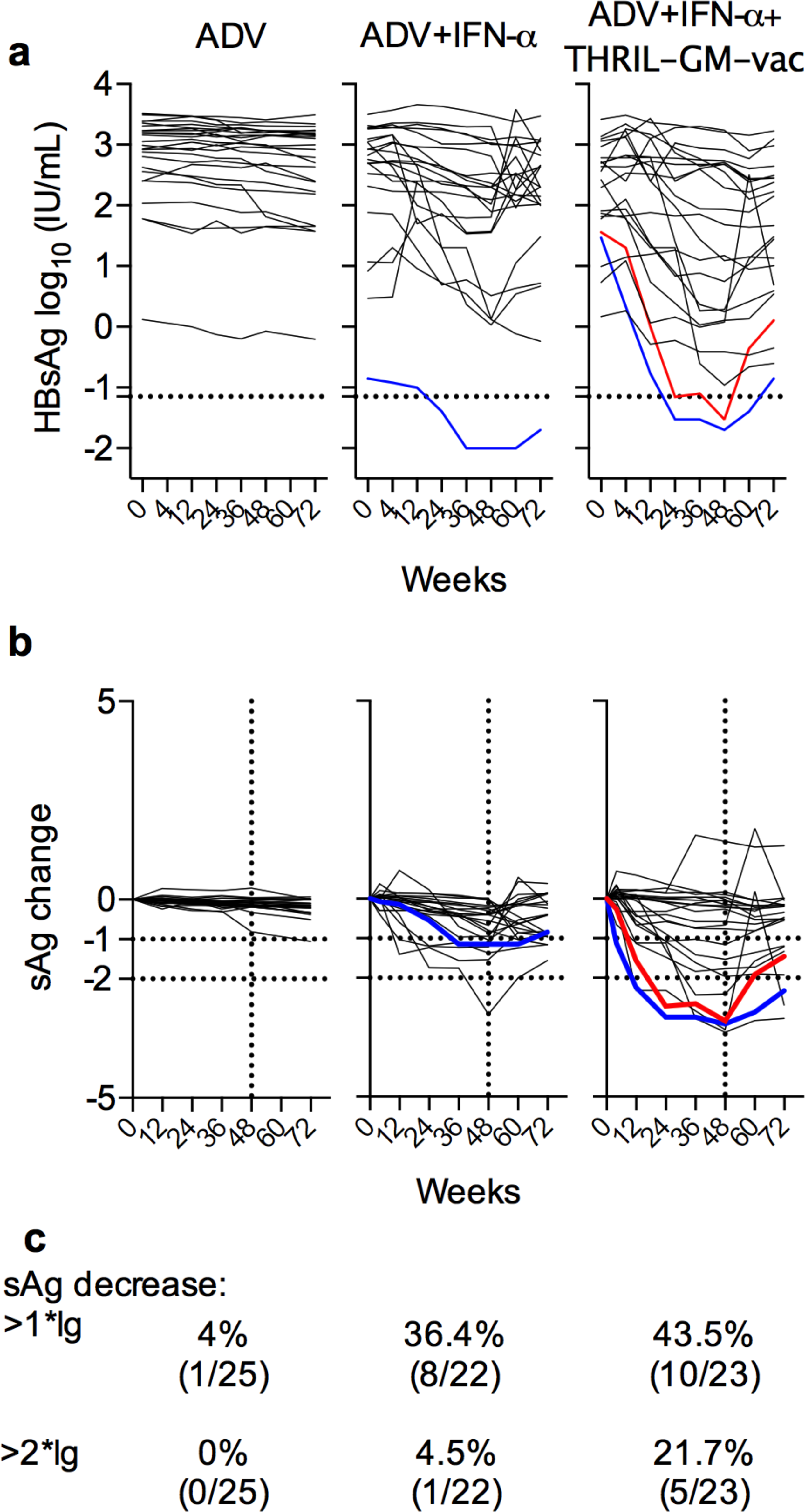
Analysis of circulating HBsAg levels before and after treatment. Serum samples were collected from each participant and analyzed using the Roche Cobas e411 platform. a) HBsAg dynamic changes: Colored lines represent patients who achieved HBsAg seroclearance (defined as HBsAg <0.07 IU/mL as indicated by the dashed line). b-c) Individual normalized HBsAg dynamics: Individual HBsAg values were normalized by subtracting the baseline level for each patient. Horizontal dashed lines represent 1-log and 2-log decreases in HBsAg (sAg). Vertical dashed lines represent the end of treatment at week 48. Solid lines represent the HBsAg dynamics for each patient.

As is shown in Figures 2b and 2c, the dynamics of the HBsAg levels were normalized by removing the HBsAg levels from week 0, which allowed for a more accurate characterization of the treatment’s effects on serum HBsAg.

Patients who had HBsAg levels that were decreased by 1-log10 or 2-log10 from their baseline levels in week 0 were classified as having had a successful treatment. In Group 1, treatment with ADV did not result in any cases of HBsAg seroclearance, since this was not an effect of the treatment. It also had very little impact on the levels of antigen in general; just one patient saw a drop in their HBsAg level of one log10, and no patients experienced a decrease in their HBsAg level of two log10 or higher (Fig. 2b, 2c). Notably, the treatment regimen consisting of IFN-α and ADV (Group 2) was successful in reducing HBsAg levels; 33.33% (8 out of 24) of patients exhibited a 1-log10 decline, and 4.17% (1 out of 24) of patients showed a 2-log10 decrease. Not surprisingly, the most significant reduction of HBsAg was observed in the presence of THRIL-GM-Vac (Group 3), where 43.5% (10 out of 23) patients showed more than a 1-log10 decrease and 21.7% (5 out of 23) showed more than a 2-log10 decrease of HBsAg level, including two patients who reached HBsAg seroclearance (Fig. 2b, 2c). In light of these data, it may be concluded that therapy with THRILGM-Vac, in conjunction with ADV and IFN-α, has the potential to enhance the frequency of HBsAg seroclearance.

### THRIL-GM-Vac breaks the immune tolerance of HBsAg

Regulatory T cells (Treg) are a significant tolerance determinant for chronic HBV infection, while HBsAg is the primary tolerance inducer. Both of these factors are important for tolerance development. In the past, it was shown that administration of THRIL-GM-Vac to HBV-transgenic and AAV-HBV-infected mouse models will result in an immune response profile characterized by a down-regulation of Treg cells. Throughout this study, HBsAg-specific CD4+FoxP3+ Treg cells were analyzed by a flow cytometry method from the patient’s peripheral blood mononuclear cells (PBMC) and evaluated both during the therapy and during the follow-up period (SupFig. 1). The HBsAg-specific Treg cells were not significantly affected by the ADV-only therapy (Group 1) (Fig. 3a), however, the numbers of Treg cells did decrease under the ADV+IFN-α combination treatment (Group 2), albeit not to a statistically significant change (Fig. 3b). Treg cells that were antigen-specific dropped by a substantial amount in Group 3, which received therapy with THRIL-GM-Vac (Fig. 3d, SupFig. 1b).

**Figure 3.**
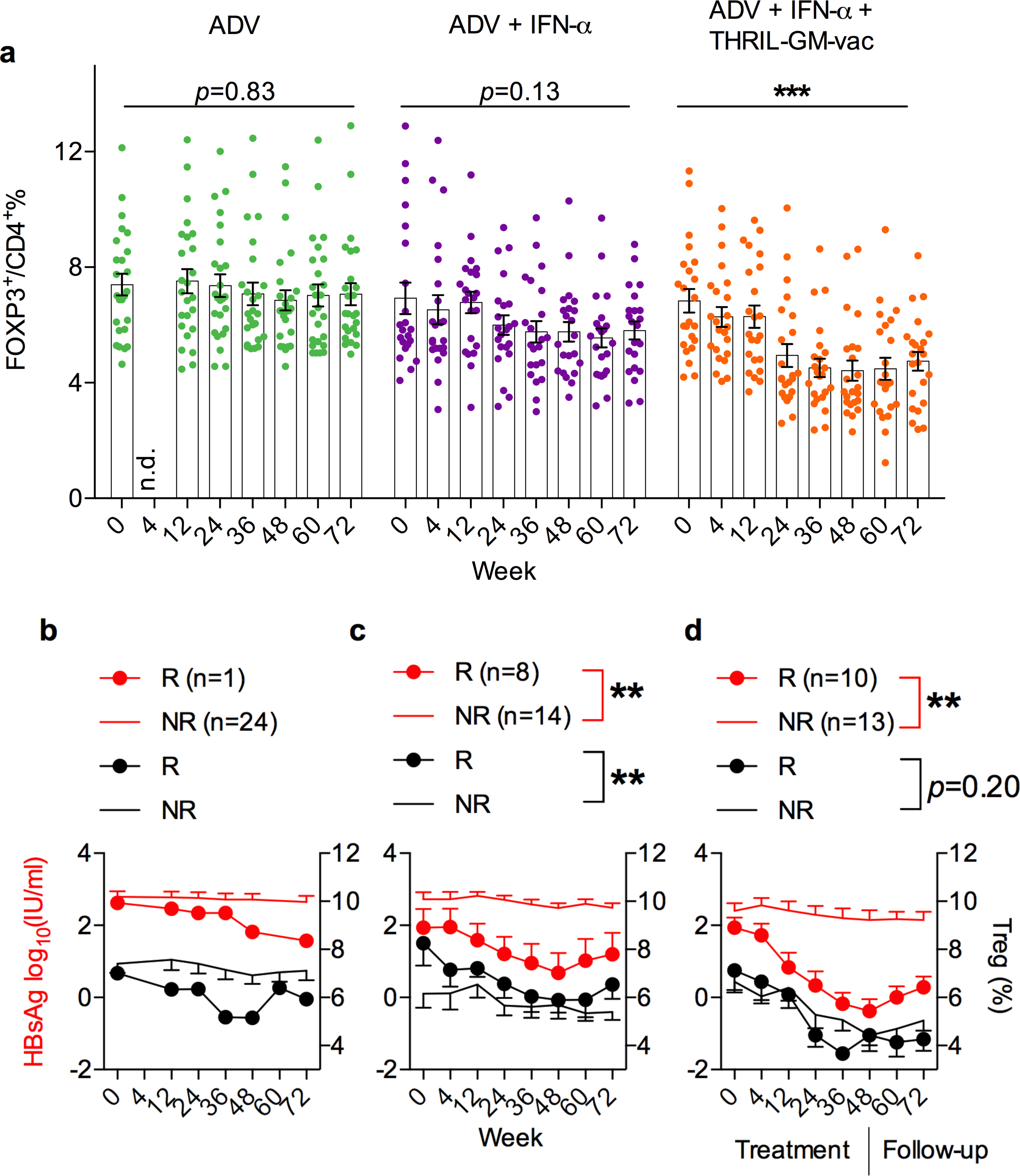
Treg cell dynamic changes throughout treatment. Peripheral blood mononuclear cells (PBMCs) were isolated from all patients and stimulated in vitro with a set of HBsAg peptide pools before analysis by flow cytometry. a) Changes in HBsAg-specific CD4+Foxp3+ Treg cells: This panel shows the percentage changes in HBsAg-specific CD4+Foxp3+ Treg cells in patients’ PBMCs throughout treatment. Each dot represents an individual patient. ***, represents p< 0.005 compared by wk 72 vs wk0. b-d) Correlation between HBsAg and Treg: These panels show the correlation between HBsAg levels and Treg percentages in group 1 (b), group 2 (c), and group 3 (d). The red line represents the HBsAg level, the black line represents the Treg percentage, and the dotted line represents responders (R). Plain lines represent non-responders (NR). P-values are indicated by **, p < 0.01.

We began by defining patients as either "responders" or "non-responders" to have a better understanding of the significant changes in HBsAg level that occurred as a result of the three distinct therapies and how those changes were related to changes in Foxp3+Treg. Responders were defined as patients whose HBsAg levels decreased by more than one log10, while "non-responders" were defined as patients whose HBsAg levels remained the same after treatment (a reduction of one log10). Figures 3b–3d illustrate the association between the subjects’ levels of HBsAg and the changes in antigen-specific Treg levels that occurred over time. After receiving therapy, "responders" had lower levels of HBsAg, which is linked with lower levels of Treg cells. This suggests that the loss of HBsAg may be a result of breaking immunological tolerance. When compared to responders, non-responders showed much lower declines in Tregs, which is consistent with the notion that the reduction in Tregs is coupled with the decrease in HBsAg. In addition, there was a decline in the total number of T-reg cells seen in non-responders who were in Group 3. (Fig. 3d), which suggests that THRIL-GM-Vac is robust enough to disrupt the Treg-mediated tolerance in each patient, even though other variables may potentially contribute to the clearance of HBsAg in this group.

Because of HBV tolerance, a person may have a reduction in their antiviral immunity, which includes a reduction in the frequencies of CD4+ helper T cells and CD8+ cytotoxic T cells [21]. We investigated whether or if the recent breakdown of HBV tolerance and the related decrease in Treg were reciprocally paralleled by increasing levels of HBsAg epitope-specific CD4+ and CD8+ Teff cell activation. The effector-cell populations were characterized according to Figure 4a and SupFigures 2a and 3a. The notion that immunological activation occurred after the tolerance break is supported by the fact that the IFN-γ response was considerably activated in Group 3, in both CD4+ T cells (Fig. 4b, SupFig. 2b) and CD8+ T cells (Fig. 4c, SupFig. 3b).

**Figure 4.**
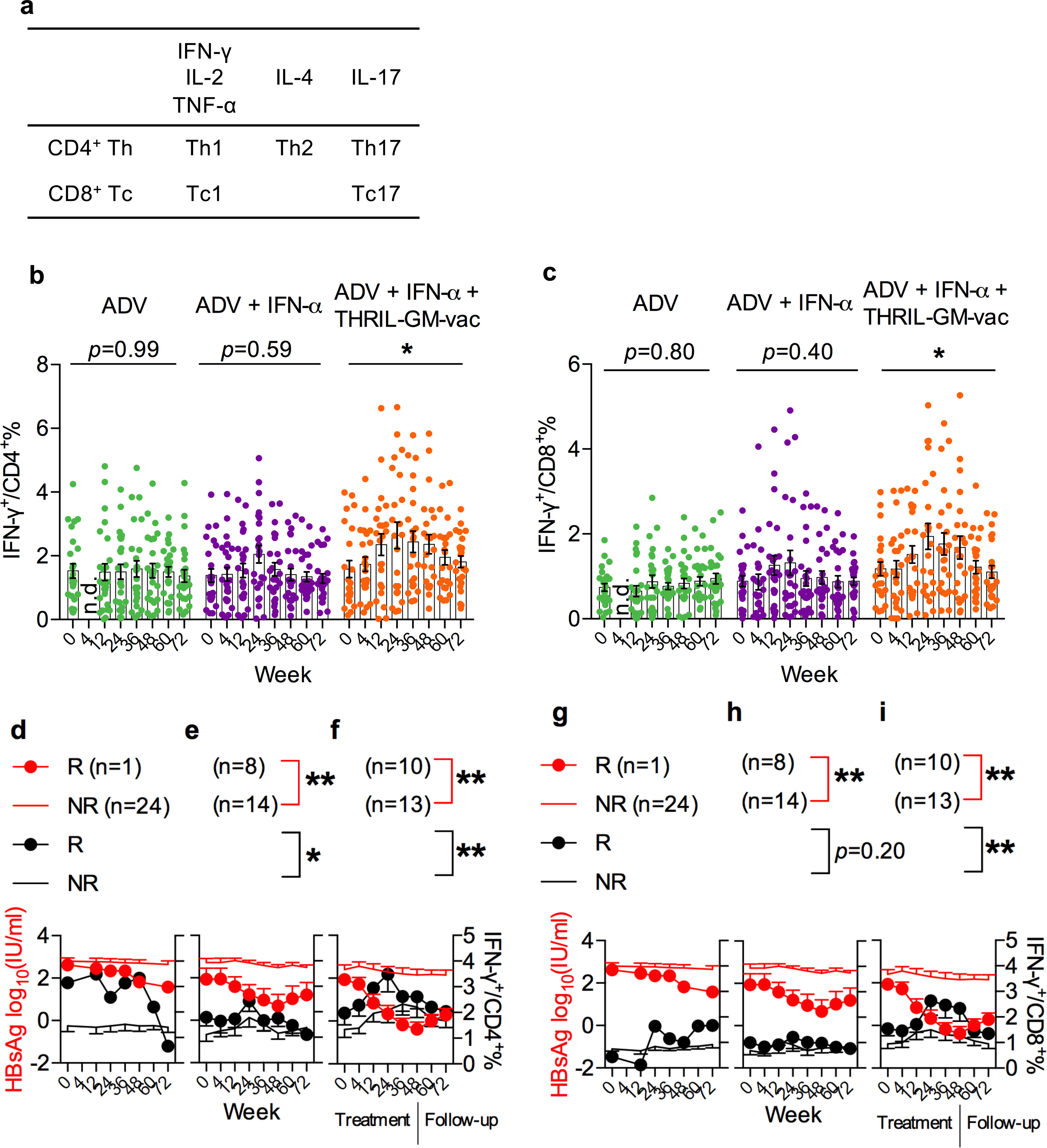
Correlation of Helper CD4 T cells and cytotoxic CD8 T cells in HBsAg serocleanrance. Flow cytometry analysis gated on HBsAg peptides stimulated CD4 and CD8 T cells with intracellular staining for IL-2, TNF-a, IL-17, IL-4, or IFN-γ, respectively. a) cytokines tested and indicated T cell subsets. b) Percentage of Th1 (IFN-γ^+^ in CD4 T cells) in total CD4 T cells, c) Tc1 (IFN-γ^+^ in CD8 T cells) in total CD 8 T cells. Dots represent each patient. d-f) Th1 negatively correlated with HBsAg (sAg) in responders. g-i) Tc1 negatively correlated with HBsAg (sAg) in responders. ** indicates p<0.01.

In Figures 4d-4i, the data on IFN-γ and antigen from responders and non-responders are plotted side by side to illustrate correlations between HBsAg decrease and anti-viral cytokine dynamics. This patient’s anti-viral immune responses, rather than the anti-viral drug action, may have affected viral replication during the ADV treatments; perhaps the patient was a spontaneous responder’ rather than a drug responder. There was only one responder in the ADV-treatment group (Group 1), and this patient had more robust Th1 and Tc1 responses than the other patients in the group (Figs. 4d and 4g). The responders in Group 2 had more robust Th1 and Tc1 responses (Figs. 4e and 4h), but antigen clearance was not substantially different when compared with the non-responders. Responders in Group 3 had up-regulation of Th1 even in the early period (week 4 to week 24), followed by sequential activation of Tc1 from week 12 through the end of treatment at week 48. These findings revealed significant correlations between the decline in HBsAg and the robust Th1 and Tc1 responses. During this time, the levels of HBsAg decreased the most noticeably from week 4 to the completion of the therapy. Under the influence of THRIL-GM-Vac, these substantial correlations revealed that anti-viral CMI, which was mediated by robust activation of both Th1 and Tc1 cells, was responsible for the clearance of HBsAg.

In the context of CHB immunotherapeutic treatments, the possibility of causing damage to the liver is a key point of concern about possible adverse effects. As a result, we additionally examined any dynamic variations in ALT levels that occurred in each patient over the course of the trial. As can be seen in Figure 5, ADV therapies had no effect on ALT levels, which reflects their inability to activate the immune system. After therapy, the ALT levels of several patients in the ADV+IFN group (Group 2) increased to high but acceptable levels, suggesting that they had minor liver injury. It seems that immunotherapy with THRIL-GM-Vac is responsible for raising the ALT levels in Group 3, as shown by an almost twofold increase in average ALT levels at week 24 followed by a subsequent reduction in those levels. This may be a reflection of a positive benefit caused by the targeted clearance of infected HBV hepatocytes by antigen-specific immune destruction.

**Figure 5.**
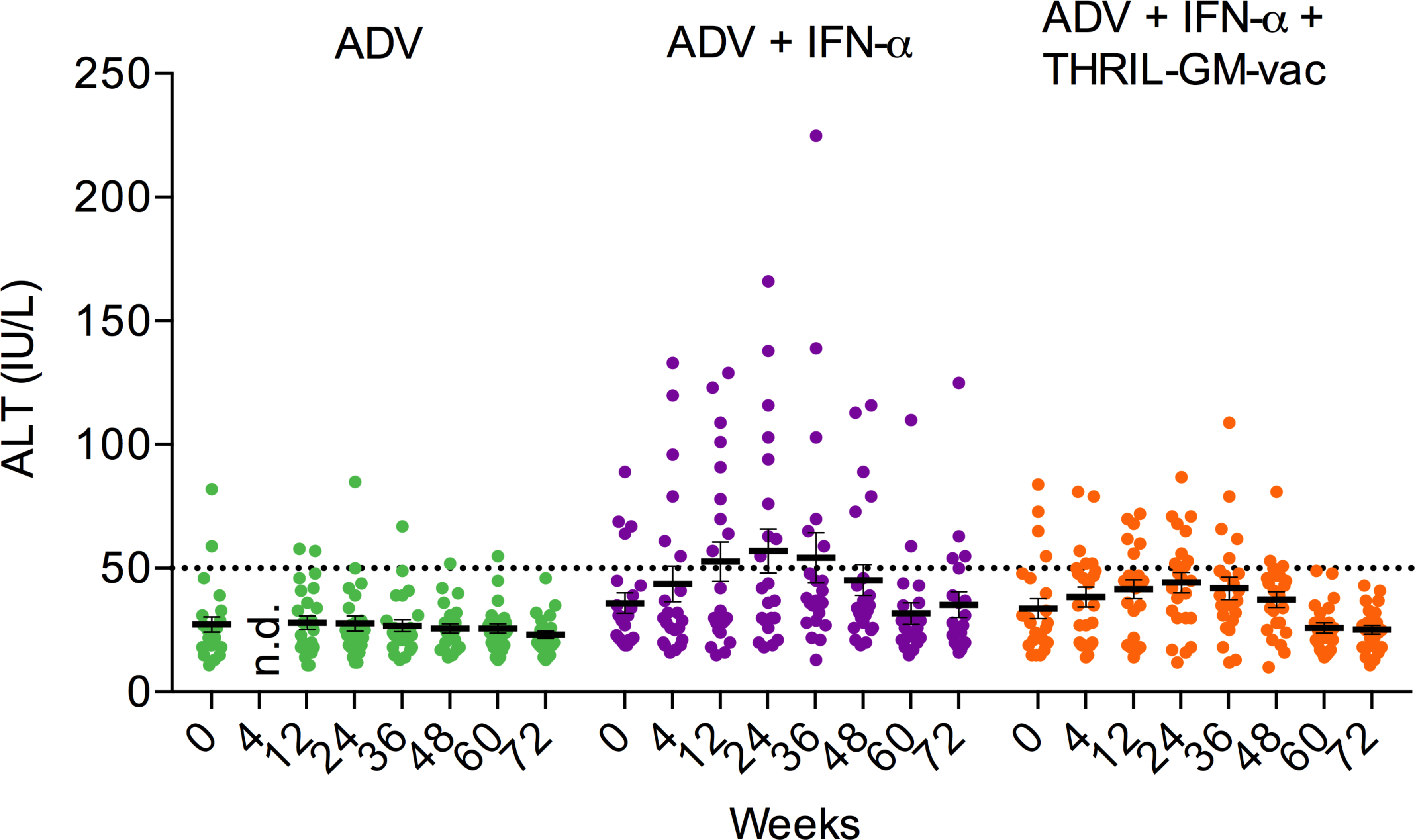
ALT dynamics over the treatment period in each patient. The dashed line represents the normal ALT level. Each dot represents each individual. There were no statistically significant differences among the groups.

Together, our findings imply that effective HBsAg clearance by supplementary THRIL-GM-Vac therapy correlates with the breakdown of tolerance by activating antigen-specific cell-mediated immunity to eliminate infected cells and that this results in minimal liver damage.

### THRIL-GM-Vac favored HBsAg clearance in patients with medium-to-low baseline antigen levels

Our clinical experiences have led us to believe that the amount of HBsAg that was present at the beginning of therapy may determine how well it is administered. For instance, individuals who had already achieved low HBsAg baseline levels thanks to the administration of NA therapies did not benefit from IFN-α therapy. This connection was also seen in the patients of Group 2 who were given the ADV+IFN-α treatment in this investigation. As a result, we exhibited the association between the baseline levels of HBsAg and the maximum values of HBsAg change, Treg change, Th1 change, and Th2 change over the treatments (Fig. 6).

**Figure 6.**
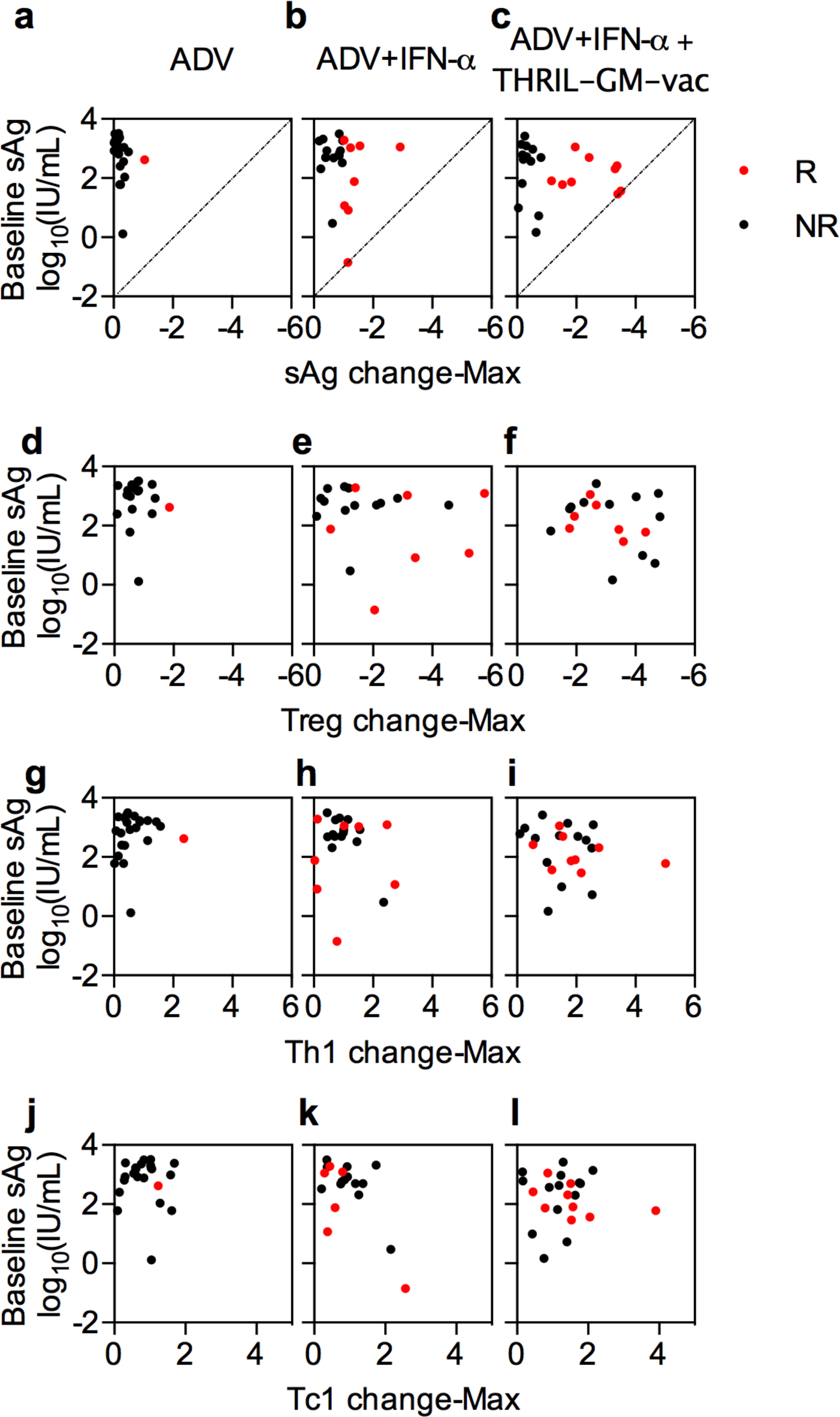
Relationship of each patient’s HBsAg baseline with HBsAg reductions (a-c) by wk48 the end of the treatments, and with each immuno-profile on the Treg change (d-f), Th1 change (g-i), and Th2 change (j-l) during treatment to the baseline levels of HBsAg. the non-responders (NR) were indicated as black dots, and the responders (R) as red dots.

The non-responders tend to cluster in the upper-left corner of each panel, which is consistent with an association between a higher antigen load and a lower reduction of antigen load in response to treatments. This can be seen in Figures 6a to 6c, which plot the maximum changes in HBsAg levels against the baseline levels. The responders are distributed near the diagonal HBsAg clearance line (HBsAg 0.07 IU/mL). The ADV+IFN-α group (Group 2; Fig. 6b) includes just one patient with seroclearance, and this patient’s baseline level is somewhat low at 0.14 IU/mL; the patients with larger baselines exhibit modest antigen reduction responses. However, when the THRIL-GM-Vac was added to the therapies (Group 3), two patients with medium-to-low baseline HBsAg obtained seroclearance (36.36 IU/mL and 29.32 IU/mL), whereas patients with higher baseline exhibited greater HBsAg decrease towards the clearance line (Fig. 6c).

Increased levels of circulating HBsAg are often the cause of more acute immunological exhaustion and impairments of anti-viral immune potential. Effective therapy has been demonstrated to reduce Treg and up-regulate Th1 and Tc1 responses, particularly in individuals with medium-to-low baseline HBsAg levels (baseline levels ranging from 10 to 1000 IU/mL) [22]. We plotted the number of Tregs that were reduced and the number of anti-HBV cytokines that were increased against the amount of antigen that was present at the beginning of the study. This allowed us to determine whether or not THRIL-GM-Vac could reverse immune impairment even in patients whose baseline HBsAg levels were higher. When compared to the ADV therapy on its own, the addition of IFN-α to the ADV treatment (Group 2) resulted in a modest increase in the percentage of patients who had substantial alterations in their immune responses. This was seen in Figures 6e-6k. However, when the THRIL-GM-Vac was added to the ADV+IFN-α therapy regime, the majority of patients in Group 3 demonstrated large improvements in cellular immune responses. This was the case regardless of the volume of the antigen load at the beginning of the study (Fig. 6f-6l).

Breaking immunotolerance in CHB patients using immunotherapeutic treatments with the THRIL-GM-Vac, which consists of three treatments with GM-CSF followed by an administration of a recombinant HBV vaccine, is effective in achieving HBsAg seronegativity, also known as a cure. These results, when taken together, show that HBsAg seronegativity can be achieved. Importantly, when combined with a current conventional approach that includes anti-viral medication ADV and IFN-α therapies, this innovative THRIL-GM-Vac therapy was effective. The host cellular immune responses that were evaluated in this investigation provided support for the conclusion that the down-regulation of tolerogenic Treg cells and the substantial amplification of HBsAg-specific immune responses were probably the major mechanisms responsible for the increased therapeutic efficacy.

## DISCUSSION AND CONCLUSION

In pursuit of an HBV functional cure, here we present our first clinical effectiveness results of a new GM-CSF-based intervention of chronic HBV patients (CHB). Cohort 3 treated with THRIL-GM-Vac therapies achieved significant HBsAg reductions (2log10) in 21.7% of participants, compared to 0% in cohort 1 and 4.17% in cohort 2. Furthermore, a strong correlation was identified between the degree of HBsAg reduction and antigen-specific cell-mediated immunity. The primary findings are detailed as follows:

First, significant reductions in the levels of HBsAg were seen in the patients in Group 3 who had been treated with THRIL-GM-Vac immunotherapy (Fig. 2). During the 48 weeks of immunotherapy, there was a significant reduction in the number of HBsAg-specific Treg cells in the ’responders’, as well as a significant increase in anti-HBV CMI, particularly in the ’responders’. This was evidence that immunotolerance induced by HBsAg was destroyed, and it was demonstrated by both of these changes (Figs. 3 and 4). Although the antigen-specific CMI began to decrease after week 48, the major mobilization of CMI might be able to be prolonged with a strategy that employs immunotherapeutic treatment for a longer period and makes use of the ratio of regulatory T cells to effector T cells as an efficacy biomarker.

Second, when the immunotherapeutic treatments had been completed at week 48, HBsAg levels slowly increased again, although Tregs remained at low levels throughout the study (Figs. 2 and 3). This trend was closely linked to declining levels of Tc1 and Th1 cells in responders (Figs. 3 and 4). The reduction in CMI should be investigated further in future clinical studies that either last for longer and include more vaccination courses or have more vaccine doses administered within the same time frame (48 weeks). In the meantime, the therapy might be improved by the creation of novel vaccination strategies. For instance, a sequential treatment schedule with a novel kind of NA therapy may inhibit viral replication more effectively before the THRIL-GM-Vac therapies are initiated. This, in turn, may produce even larger numbers of antigen specific CD4+ Th1 cells and more efficient CD8+ Tc1 cells.

Third, we found that not all CHB patients reacted well to the THRIL-GM-Vac treatments, even though some of them were considered "responders". This points to the fact that patients’ immunological responses to the therapies, often known as their immune sensitivity, were very different from one another. These distinctions might be predetermined, and an assessment of the patient’s immunological profile at baseline could yield surrogate markers that indicate whether the patient is suitable for this treatment approach. The findings suggested that the baseline immune profile is a significant parameter to predict the outcome and that it might be used to pre-screen patients with CHB who were "sensitive" to determine whether they would benefit from the treatment.

Fourth, our results are in line with the theory that subsequent generations of Th1 cells and Tc1 cells are necessary for HBsAg decrease and clearance in responders, and this concept is supported by our research. This may lead to the creation of a generalized immune-surrogate endpoint if it can be shown that this is the case.

While our GM-CSF-based strategy demonstrated significant improvement over frontline NA or IFN-α treatments, we identified limitations, particularly in patients deviating from our previous animal model results. Firstly, THRIL-GM-Vac therapy displayed the greatest success in patients with medium-to-low baseline HBsAg levels (10-100 IU/ml) compared to those with high levels (>1000 IU/ml). This finding, coupled with the therapy’s safety profile in terms of ALT preservation, suggests promising avenues for future iterations aiming to enhance immune activation potency. Secondly, despite achieving nearly 100% functional cure in mouse models, a clear disparity in effectiveness emerged in this human trial (8.7%) [19]. This reinforces the well-established notion that animal models often provide starting points rather than definitive guides for human diseases. We recognize that the complexities of patients, especially those with advanced CHB, significantly differ from those observed in controlled animal settings. However, we remain steadfast in our conviction, backed by years of dedicated research, that further protocol refinements hold the potential to significantly improve therapeutic impact for a broader patient population, including those with higher baseline HBsAg levels.

## MATERIALS AND METHODS

### Drugs

Commercial products, Adefovir Dipivoxil Tablets at 10mg/tablet were manufactural produced under GMP compliant conditions and donated by Cisen Pharmaceutical Co., Ltd (Jinan, Shandong, China), Recombinant Human Interferon-α2b Injection at 5mIU/injection by Beijing Kawin Technology and human recombinant GM-CSF (Topleucon) at 75ul/dose were manufactural produced under GMP compliant conditions in *E. Coli* and donated by Amoytop Biotech (Xiamen, Fujiang, China). Human recombinant HBV vaccines 20ug/dose were manufactured and produced under GMP-compliant conditions in the yeast system and donated to the study by Kantai Biologicals (Shenzhen, China).

### Ethics statement

This clinical trial was registered in the Chinese Clinical Trial Registry (ChiCTR-TRC-13003254) as a multi-centered and randomized. However, we used frozen PBMCs of three cohorts of patients from one of ten participating hospitals. This study was approved by the Research Ethics Committee of Huashan Hospital, Fudan University. Each participant in the study was enrolled in Huashan Hospital, Fudan University after being given an informed written consent form and signed.

### Patient enrollment and treatment schedules

All recruited CHB patients had been previously treated with anti-viral NA therapy and had reached a status of HBeAg-negative while remaining HBsAg-positive(HBsAg<5000IU/ml). Criteria for recruitment also included HBV DNA level (<500 copies/mL), normal blood alanine-aminotransferase test, and consolidation therapy for more than one year. Patients were not enrolled for the study if they were pregnant or had concomitant human immunodeficiency virus infection, hepatitis C or D virus, primary biliary cholangitis, autoimmune hepatitis, Wilson’s disease, alcoholic or non-alcoholic steatohepatitis, or history of liver cirrhosis or hepatocellular carcinoma(HCC). Patients were randomly assigned into three groups: Group 1, adefovir NA treatment (ADV); Group 2, ADV plus IFN-α treatment (ADV+IFN-α); Group 3, ADV+IFN-α plus THRIL-GM-Vac treatment (ADV+IFN-α+THRIL-GM-vac).

ADV was administered orally every day (QD), IFN-α was administered as an intramuscular (i.m.) injection on alternate days (QOD), and THRIL-GM-Vac was administered as daily (QD) subcutaneous (s.c.) injections of GM-CSF for three consecutive days followed by an i.m. injection with Human HBV Vaccine at the same site on the fourth day on weeks 0, 4, 12, 24, 36, and 48, as shown in Figure 1

### Biochemical, serological HBV analysis

Serum alanine aminotransferase activity (ALT) was determined by a fully automatic biochemical analyzer (Hitachi 7600, Tokyo, Japan). Serum HBsAg, HBsAb, and HBeAg were measured using enzyme immunoassay equipment (Cobas e411, Roche Diagnostics, Mannheim, Germany).

### Virological studies

Serum HBV DNA, HBeAg seroconversion, and the levels of serum HBsAg and ALT were monitored by an independent third-party laboratory (Clinical Laboratory, Ruijin Hospital, Shanghai, China), using the same lots of reagents. Sequential samples from one patient were tested on the same day. Abbott EIA AxSYM (Abbott, Abbott Park, IL, USA) was employed for the detection of HBsAg, HBeAg, and anti-HBe. HBV DNA was quantified by fluorescent PCR assay (PiJi, Shenzhen Co, China with a detection limit of 500 copies/mL). Architect HBsAg QT assay (Abbott, Abbott Park, IL, USA) was used for serum HBsAg quantification. Routine biochemical and hematological tests were performed at each evaluation center using automated assay systems.

### Stimulation of PBMC with HBsAg epitopes in vitro

Peripheral blood mononuclear cells (PBMC) were isolated from fresh blood by Lymphoprep^TM^ (Axis-Shield, Norway) and re-suspended in complete medium R10 (RPMI 1640 supplemented with 25 mM HEPES, 2 mM L-glutamine, 100 U/mL penicillin, 100 μg/mL of streptomycin, and 10% fetal calf serum, all from Gibco (Life technology, USA). For T cell expansion in vitro, PBMC was suspended in R10 in the presence of anti-CD3 (0.1 μg/mL; Miltenyi Biotec, USA) and anti-CD28 (0.05 μg/mL, Miltenyi Biotec) at a concentration of 5 × 10^6^ cells/mL and seeded at 1 mL/well in a 12-well plate. The immunological assays were performed on day 3 of the expansion. Antigen-specific stimulation, expanded PBMC were washed once with R10 and then stimulated with HBV peptide pools (10 μg/mL, listed in Supplementary Table 1) or medium alone (control) for 8 hours in the presence of anti-CD28 (0.1 μg/mL; Miltenyi Biotec) and brefeldin A (BD Biosciences). Cells incubated with anti-CD3 (1 μg/mL) and anti-CD28 (100 ng/mL) were used as positive controls. The plates were incubated at 37°C in a humidified atmosphere with 5% CO2 for 72 h before the cells were labeled with specific monoclonal antibodies and subjected to flow cytometry analysis.

### Flow cytometry detection

Cells after washing were stained with anti-CD4 and anti-CD8 monoclonal antibodies for 30 min at 4°C, then fixed and permeabilized using 4% paraformaldehyde (PFA, Sinopharm Chemical Reagent Co., Ltd, China) and 0.2% Triton X-100 (Genview, China). For intracellular cytokine staining, the cells were further stained with the selected fluorescent-labeled anti-human monoclonal antibodies used for flow cytometric analysis: CD8 (SK1), CD4 (OKT4), FoxP3 (PCH101), IFN-γ (4S.B3), IL-2 (MQ1-17H12) from eBioscience, TNF-α (Mab11), IL-4 (MP4-25D2), IL-17 (BL168) from BioLegend. In cytokine detection, PMA (100 ng/mL) and ionomycin (1 mg/mL) were used as positive control stimulants. An LSRFortessa (BD Biosciences) was used for flow cytometry analyses.

### Statistics

Categorical variables were described using percentages. Parametric continuous variables were presented as means ± Standard Error of Mean (SEM), while non-parametric continuous variables were expressed as the median (IQR). For continuous variables, the Mann-Whitney U test was used for abnormal distribution, the two-tailed Student’s t-test for normal distribution in two-group comparisons, and one-way ANOVA for three-group comparisons. A P value of < 0.05 was deemed significant for all analyses.

## Data Availability

All data produced in the present study are available upon reasonable request to the authors

## ACKNOWLEDGMENTS

This work was partly supported by awards of the National Major Science and Technology Research Projects for the Control and Prevention of Major Infectious Diseases in China (2017ZX10202202, 2013ZX10002001, and 2013ZX10002001003) to Drs. JM Zhang, HY Jia, LD Yang, and B.Wang, and the National Natural Science Foundation of China (81871640, 82172255) to B. Wang. We thank Dr. Douglas Lowrie for his careful editing and proofreads and Dr. Yiwei Zhong at Shanghai Medical College of Fudan University for her technical assistance.

## AUTHOR CONTRIBUTIONS

BW, JMZ, HYJ, and LDY designed the clinical study and analyzed the results. JMZ, HYJ, and LDY managed the operation of trials. FFY, HC, and LY managed patient screenings, samplings, recordings, and clinical analysis. SG, GZ, WDZ, SRZ, XYZ, CFL, FY, XJ, XZ, and SJZ performed CMI experiments and analyzed data. SG, BW, and JMZ wrote and edited the manuscript.

## CONFLICT OF INTEREST

Authors declare that they have no competing interests.

## Notes

### Competing Interest Statement

The authors have declared no competing interest.

### Clinical Trial

ChiCTR-TRC-13003254

### Author Declarations

The study was approved by the Research Ethics Committee of Huashan hospital, Fudan University. All the study participants were enrolled in Huashan Hospital, Fudan University, and provided written informed consent.

### Summary of Updates

Add reagents with sources and detailed experimental procedures in Materials & Methods; revised the abstract and discussion; add supplementary figures and a table; proofread the manuscript for accuracy and clarification. Change the order of authors' list.

