## Supplementary Table 1 for "Seroclearance of HBsAg in Chronic Hepatitis B Patients After a Tolerance Breaking Immuno-Therapy: GM-CSF, followed by A Recombinant HBV Vaccine"

**Supplementary Table 1**. HBsAg Specific Peptide Pools for T Cell In Vitro Stimulation

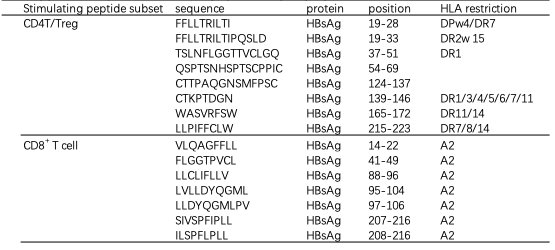

Notes:

- MHC binding predictions and epitope references: Utilize online resources like IEDB to identify potential T cell epitopes within HBsAg and assess their binding affinity to MHC molecules.
- Tailor peptide pools to specific T cell stimulations by considering available sequence information and MHC restriction of target cells.
