## Supplementary Figures 1-3 for "Seroclearance of HBsAg in Chronic Hepatitis B Patients After a Tolerance Breaking Immuno-Therapy: GM-CSF, followed by A Recombinant HBV Vaccine"

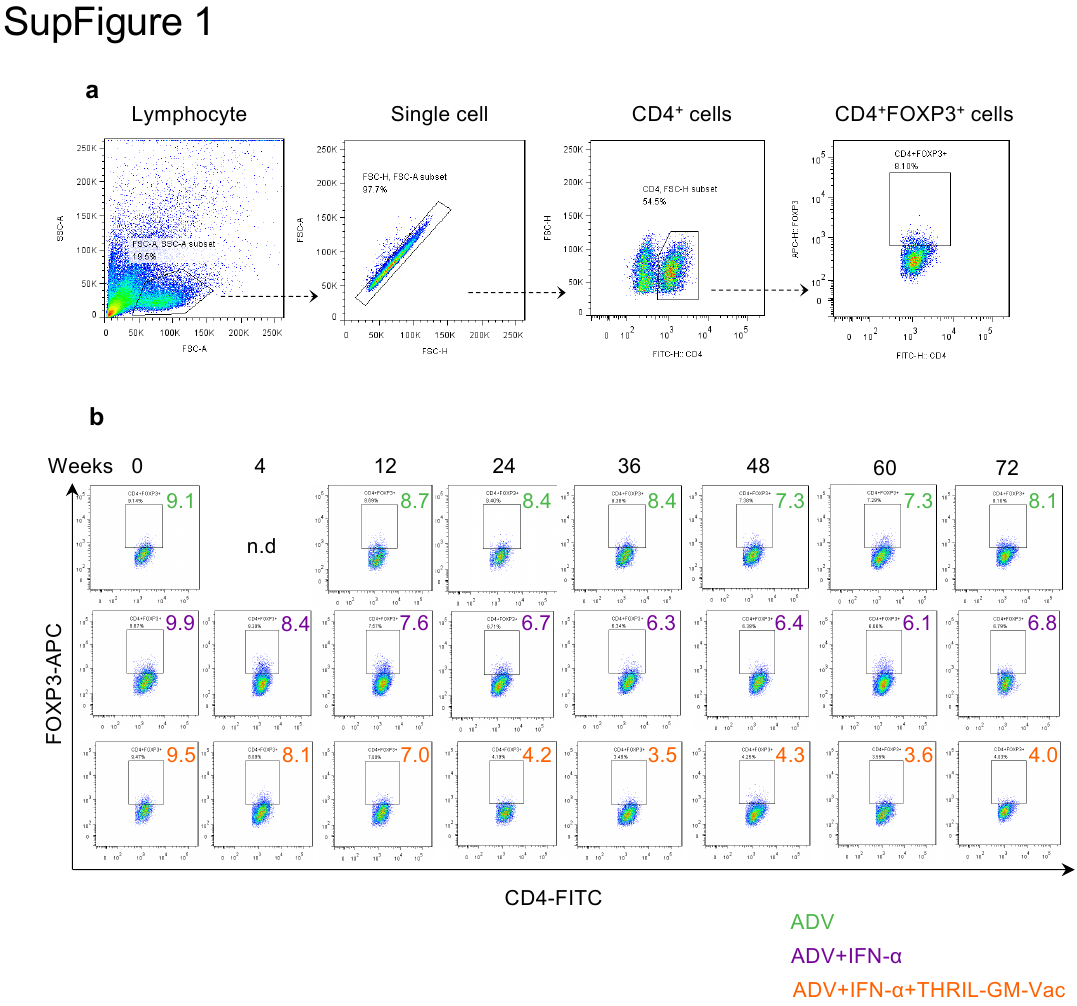
**SupFigure 1.** Gating strategy for Treg analysis and representative dot plots over the intervention course (weeks 0 to 72).

(a) Gating strategy: Lymphocytes, single cells, CD4+ T cells, and Treg cells (CD4+FOXP3+) were sequentially gated for analysis.

(b) Representative dot plots: Treg dynamics for one patient from each group are shown in a time series (each column).

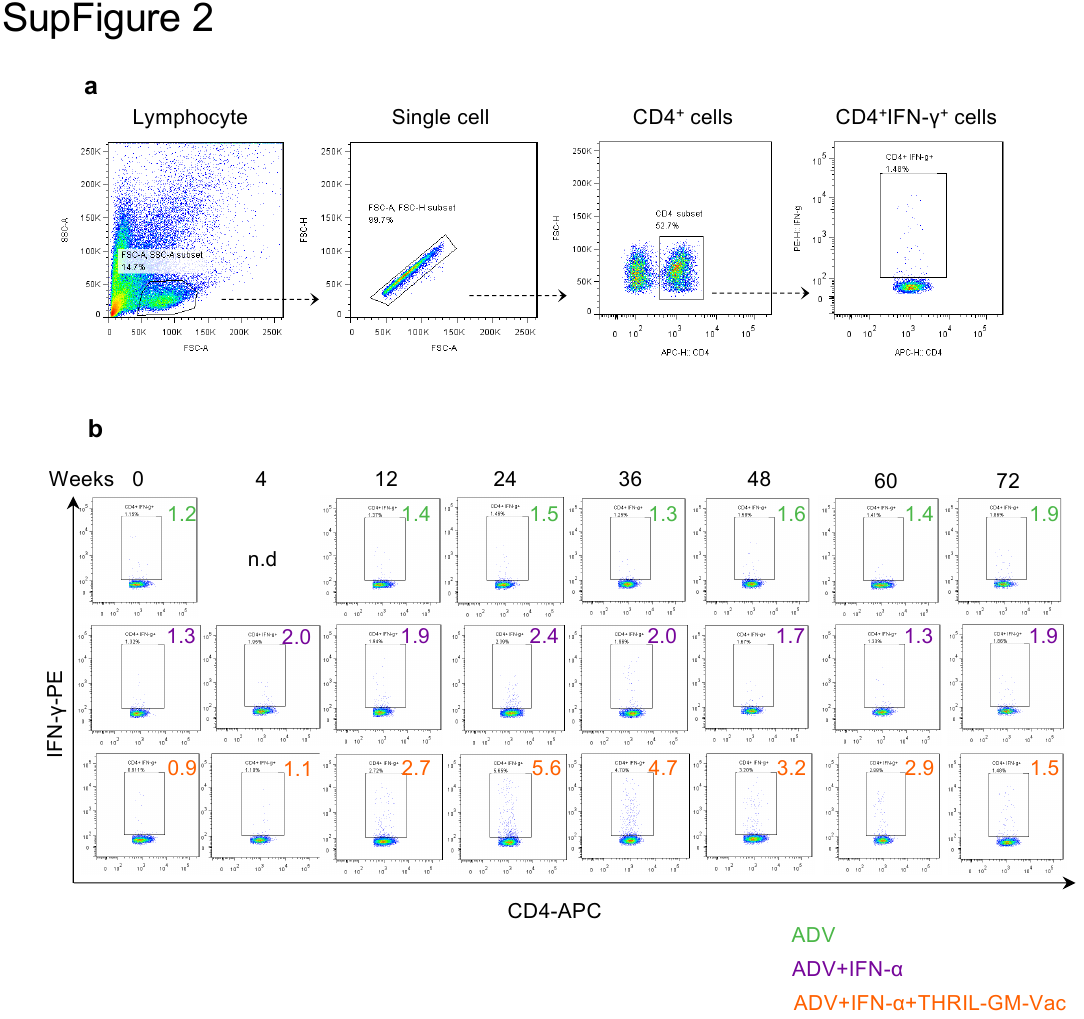
**SupFigure 2.** Gating strategy for CD4+ effector T cell analysis and representative dot plots over the intervention course (weeks 0 to 72).

(a) Gating strategy: Lymphocytes, single cells, CD4+ T cells, and Teff cells (CD4+IFN-γ+) were sequentially gated for analysis.

(b) Representative dot plots: Teff dynamics for one patient from each group are shown in a time series (each column).

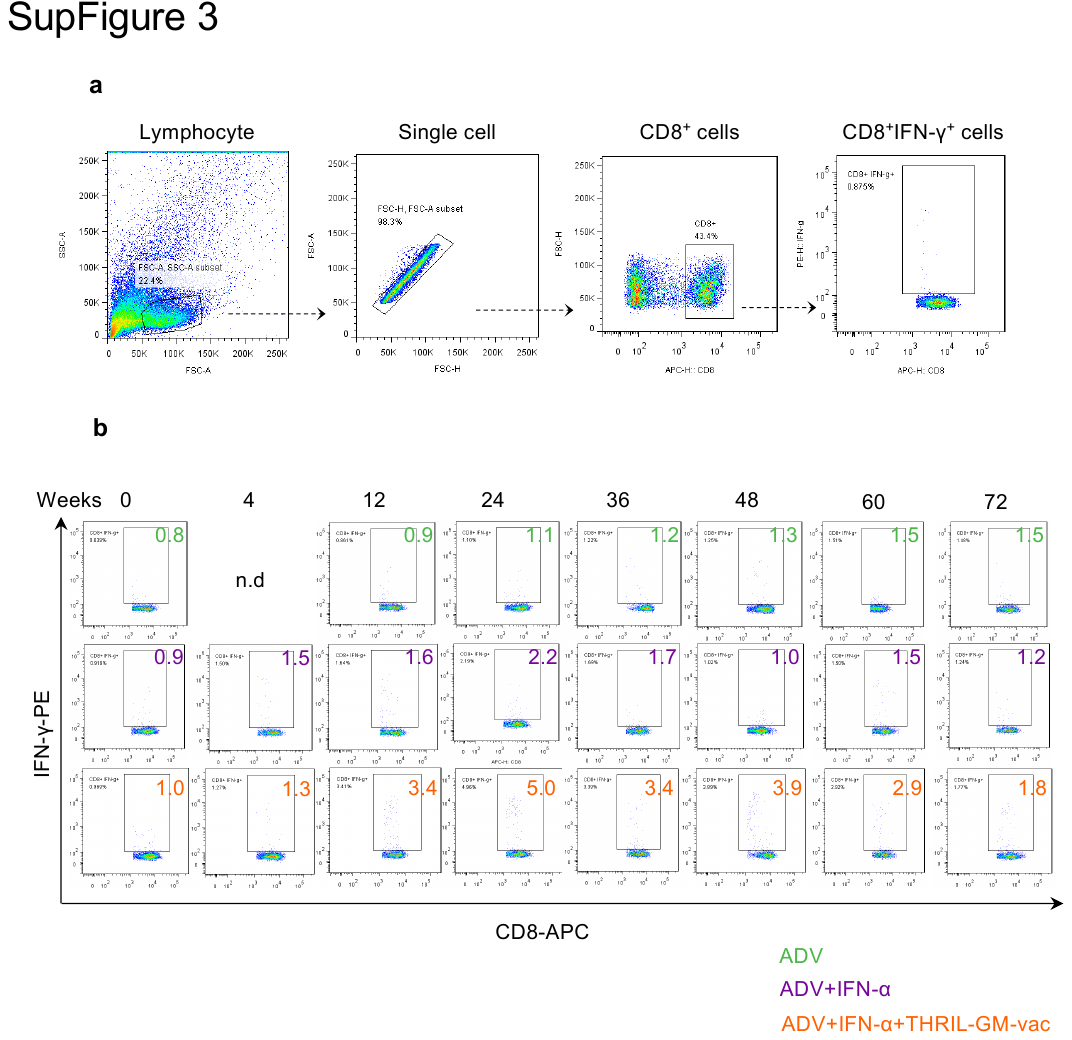
**SupFigure 3.** Gating strategy for CD8+ effector T cell analysis and representative dot plots over the intervention course (weeks 0 to 72).

(a) Gating strategy: Lymphocytes, single cells, CD8+ T cells, and Teff cells (CD8+IFN-γ+) were sequentially gated for analysis.

(b) Representative dot plots: Teff dynamics for one patient from each group are shown in a time series (each column).
